# Host biomarkers to predict the severity of acute febrile illness: A scoping review

**DOI:** 10.1101/2019.12.21.19014753

**Authors:** Matthew L. Robinson, Meklit Workneh, Sabine Dittrich, Spruha Kurlekar, Rebecca Yee, Maya C. Nirmalraj, Karen A. Robinson, Yukari C. Manabe

## Abstract

**Background:** Acute febrile illness (AFI) ranges from mild to mortal, yet severity is difficult to assess. Host biomarkers may identify patients with AFI who require a higher level of care; choosing appropriate biomarkers for this role among an expanding pool of candidates is challenging. We performed a scoping review to evaluate the performance of novel host biomarkers to predict AFI severity.

**Methods:** PubMed was systematically searched for manuscripts published January 1, 2013 to February 10, 2018 for studies reporting the association of host biomarker levels and a measure of disease severity among patients with a suspected or diagnosed cause of AFI. Identified abstracts and full text manuscripts were reviewed for eligibility by 2 reviewers. Biomarker performance was evaluated primarily by the area under the curve (AUC) of the receiver operator characteristic to distinguish severe disease. We aggregated data describing biomarker performance by AUC using weighted mean, fixed effects meta-analyses, and random effects meta-analyses.

**Results:** Among 2,303 manuscripts identified, 281 manuscripts met criteria for analysis. Data was extracted for 278 biomarkers evaluated in 45,737 participants. Among 89 biomarkers evaluated by ≥2 studies, there were 6 biomarkers (proadrenomedullin, copeptin, pro-atrial natriuretic peptide, serum triggering receptor expressed on myeloid cells-1, chitinase-3-like protein-1, and the pediatric sepsis biomarker risk model), that showed a weighted mean AUC >0.75 (range 0.75-0.84) in >500 patients over >2 studies.

**Conclusions:** Although several biomarkers show promise in predicting AFI severity across multiple studies, their test characteristics do not suggest that they may be used alone to determine AFI prognosis.

**Summary:** A systematic review and limited metanalysis of 89 host biomarkers revealed that most individual biomarkers offer modest performance in predicting the severity of acute febrile illness; several however have performance characteristics which have shown promise in multiple studies.

## Introduction

Acute febrile illness (AFI) is among the most common medical problems in low- and middle-income countries (LMICs).[1, 2] Causes of AFI vary by geographical location, season, and year and can range from minor ailments to mortal illness.[3-9] Provision of disease-specific diagnostic tests to determine AFI etiology may not always be feasible.[10] Many patients with AFI can be treated in a community outpatient setting; others require referral to a higher level of care to prevent further complications and death.[11, 12]

Accurately predicting the prognosis of patients with AFI, even in high-income countries is challenging; clinical signs and symptoms may be difficult to recognize and do not consistently predict poor disease outcome.[13-19] In LMICs, limited epidemiologic data on disease incidence, lack of diagnostics, limited healthcare worker training, and overburdened healthcare systems pose additional challenges.[20, 21] Determining the severity of AFI to rapidly identify the patients who require a higher level of care is important particularly in settings where human resources and diagnostic tests are limited.[22-26]

Host biomarkers have shown promise as prognostic indicators of disease severity in conditions ranging from cancer to inflammatory bowel disease and tuberculosis.[27-29] Novel biomarkers have also been able to discriminate bacterial from viral infection in certain cohorts.[10, 30] C-reactive protein (CRP) has been used as a measure of disease severity, including AFI, for almost 90 years.[31] Other laboratory tests in common clinical use such as procalcitonin (PCT), white blood cell count, and liver function tests have been evaluated for their ability to predict the severity of AFI.[32-38]

There is now growing interest in using biomarkers to guide referral and management of patients with AFI in LMICs.[39, 40] CRP and PCT are already available for routine clinical use, thus the evidence base describing their ability to predict illness severity is already well described.[41-43] However, advances in highly multiplex immunoassay platforms, next-generation sequencing, proteomics, and metabolomics have sharply increased the availability of data describing the host response to AFI and the ability to predict its severity.[44-47] We performed a scoping review of recently published literature on the performance of novel host biomarkers to predict the severity of AFI with the aim to inform development of diagnostics for clinical use.

## Methods

### Search strategy

Search terms were selected to identify studies which evaluated the performance of a biomarker to classify patients at risk for severe disease or death in an analyzable population consisting entirely of patients with a syndrome known to cause AFI. A non-exhaustive list of causes of AFI in LMICs was determined by review of AFI etiology in primary papers and systematic reviews.[48-50] Tuberculosis was intentionally excluded from this list as there is already a separate evidence base describing the performance of biomarkers for tuberculosis diagnosis and prognosis.[51] Causes of AFI explicitly included in the process to select search terms included sepsis, septic shock, pneumonia, diarrhea, malaria, dengue, leptospirosis, meningitis, typhoid fever and scrub typhus. Multiple manual searches were performed to identify 32 manuscripts that met our inclusion criteria and 16 near-misses. Controlled vocabulary and text words were identified from these manuscripts. A custom-built Python script was written to maximize the ratio of correctly identified manuscripts to incorrectly identified manuscripts using the most parsimonious search terms. Briefly, individual phrases were extracted from the titles, abstracts, and Medical Subject Heading terms for all 48 manuscripts. Each phrase was ranked for its sensitivity and specificity to identify manuscripts meeting inclusion criteria. Phrases were chosen that identified all 32 manuscripts warranting inclusion, and then sequentially eliminated to create the most parsimonious search query while not missing any manuscripts warranting inclusion. Using the selected terms, a search was conducted on February 10, 2018 in PubMed as shown in Table 1.

**Table 1:** Terms included in PubMed search.

| Category | Terms |
| --- | --- |
| Biomarker | biomarkers[mh] OR protein precursors[mh] OR metabolome[mh] OR biomarker[tiab] OR biomarkers[tiab] |
| Severity | severity of illness index[mh] OR survival analysis[mh] OR prognosis[mh] OR prognostic[tiab] OR mortality[tiab] |
| Acute febrile illness | sepsis[mh] OR pneumonia[mh] OR fever[mh] OR malaria[mh] OR dengue OR typhoid fever[mh] OR leptospirosis OR diarrhea[mh] OR influenza[tiab] OR meningitis[tiab] |
| Other | English[la] NOT review[pt] NOT (animals[mh] NOT humans[mh]) |
| Each category was joined by an AND statement |  |

### Study Selection

#### Study population

Studies analyzing populations with one or multiple, suspected or diagnosed causes of AFI were included. If the analyzed study population included patients without a suspected or diagnosed cause of AFI, the study was excluded (i.e., patients with systemic inflammatory response syndrome without sepsis). Hospital-acquired infections were not considered cases of AFI and their presence in the study population was cause for study exclusion. Studies in which all participants had malignancy, neutropenia, or organ transplantation were excluded. Studies that included only participants under 6 months of age were not considered. Interventional trials were excluded. Papers included only English language papers published from January 1, 2013 to February 10, 2018.

#### Biomarkers

The studies included in this review tested one or more biomarkers produced by human hosts and used the tests to assess the severity of AFI. Studies were restricted to those reporting biomarker measurement in a routinely available body fluid - blood or its components, saliva, and urine. Biomarker measurement of bodily fluids requiring invasive sampling (e.g. cerebrospinal fluid, pleural fluid) were not considered. Biomarkers already available for routine clinical use were excluded including: complete blood count (CBC) and its components, CRP, PCT, liver function tests, kidney function tests, lactate, routine endocrine tests like thyroid function tests and cardiac biomarkers such as troponin and pro-BNP. Biomarkers reported from large-scale discovery data sets such as generated in transcriptomic or proteomic studies without subsequent validation were excluded.

#### Measure of severity

Only studies reporting disease severity in terms of association with mortality, organ dysfunction, established severe disease classification system, or clinical severity score were included. Studies that reported kidney injury as the only measure of disease severity were excluded. Studies reporting only biomarker levels collected after 48 hours of presentation to healthcare were excluded. If the investigators performed more than one biomarker measurement, only the earliest measurement was considered. If more than one measure of disease severity was reported, preference was given to mortality as an outcome for further analysis.

### Screening Process

Each abstract was reviewed for inclusion by 2 independent reviewers. When the 2 reviewers did not reach agreement, a third reviewer was sought. After abstract review, provisionally included abstracts were subject to full text review again by 2 independent reviewers. Inter-reviewer disagreement on full-text reviews was handled similarly to abstracts. Abstract and full text review were managed using Covidence.[52]

### Quality Assessment

We adapted the Quality Assessment of Diagnostic Accuracy Studies-2 (QUADAS-2)[53] into a simplified scoring system to report the risk of bias and concern for applicability using the QUADAS-2 domains of patient selection, index test, reference standard, and flow and timing. One point each was given if the study design was prospective, participants were enrolled consecutively, and biomarker cutoff values were defined prospectively. One point each was subtracted if the study employed a case control design, participants with hospital-acquired infections were not explicitly excluded, or ambiguous methods or reporting did not explicitly exclude patients without infections.

### Data Extraction

Information was extracted from individual studies into a custom Microsoft Access database. Key information extracted from each study included the level of care at the time of patient enrollment which was categorized nonexclusively as outpatient, emergency department, inpatient ward, and intensive care unit (ICU). The age groups of participants were categorized as including children under age 5 years, children between age 5 to 18 years, and adults. Non-exclusive diagnoses of the study population were recorded. Disease severity or patient outcome was recorded either categorically or in correlation with existing severity scores. Categorical patient outcome measures included death, septic shock, severe sepsis, respiratory failure, subsequent requirement for ICU admission, and severe disease according to disease-specific standards. The p-value results of significance testing of an association of a biomarker with an unfavorable patient outcome or severe disease was extracted. The strength of the association as reported by area under the curve (AUC) of the receiver operator characteristic (ROC), logistic regression odds ratio, and Cox proportional hazard ratio. Unadjusted measures of association were recorded preferentially over adjusted measures if both were reported as adjustment was performed infrequently and heterogeneously. Correlation with severity scores including Acute Physiology and Chronic Health Evaluation-II (APACHE-II),[54] Sequential Organ Failure Assessment (SOFA), CURB-65, Mortality in Emergency Department Sepsis (MEDS), Multiple Organ Dysfunction Score (MODS), Pneumonia Severity Index (PSI) was recorded by Spearman r values, p values, and by severity score above or below a defined biomarker cutoff where reported.

### Biomarker Categorization

Each reported protein biomarker was queried for its inclusion in the Uniprot database.[55] Biomarkers found to be reported by multiple names were assigned one name for further analysis. Additionally, biomarkers and their prologues or breakdown products (e.g. adrenomedullin and pro-adrenomedullin) were reported together. The function of each biomarker reported by more than one study was determined by query of Uniprot and Cytoscape as well as additional literature review.[56] A flowchart was created placing the biomarkers in their primary immunological pathway. These immunological pathways were grouped into eight immune functions to simplify analysis and interpretation. Biomarkers that had been analyzed in more than one study were grouped parsimoniously according to their immune function, where applicable: vasoactivity, proinflammatory cytokines, pathogen-associated molecular pattern (PAMP), and danger associated molecular pattern (DAMP) recognition, macrophage differentiation, leukocyte migration, innate effector mechanism, and immune regulation.

### Data Analysis

For each biomarker, summary statistics describing the available evidence base to predict the severity of AFI were compiled including number of studies, combined number of participants, diagnoses, and level of care of participants. A weighted median and mean AUC was assigned for each biomarker by calculating the median and mean AUC across studies weighted by the number of participants in each study. The most promising biomarkers were shortlisted by the following criteria: inclusion of > 500 participants in ≥ 2 studies with a weighted mean AUC of > 0.70 where ≥ 75% of studies showed an association with the outcome of interest. The most promising biomarkers were further shortlisted by restricting to biomarkers with a weighted mean >0.75. Additional sub-analyses were performed by restricting to studies performed only in LMICs that did not include participants admitted to the ICU at the time of enrollment.

## Results

### Search results

The initial search yielded 2,303 results (Figure 1). Abstract review identified 444 publications that provisionally fit inclusion criteria. Review of full text manuscripts eliminated 163 publications due to reasons identified in Figure 1, leaving 281 manuscripts for data review.

**Figure 1:**
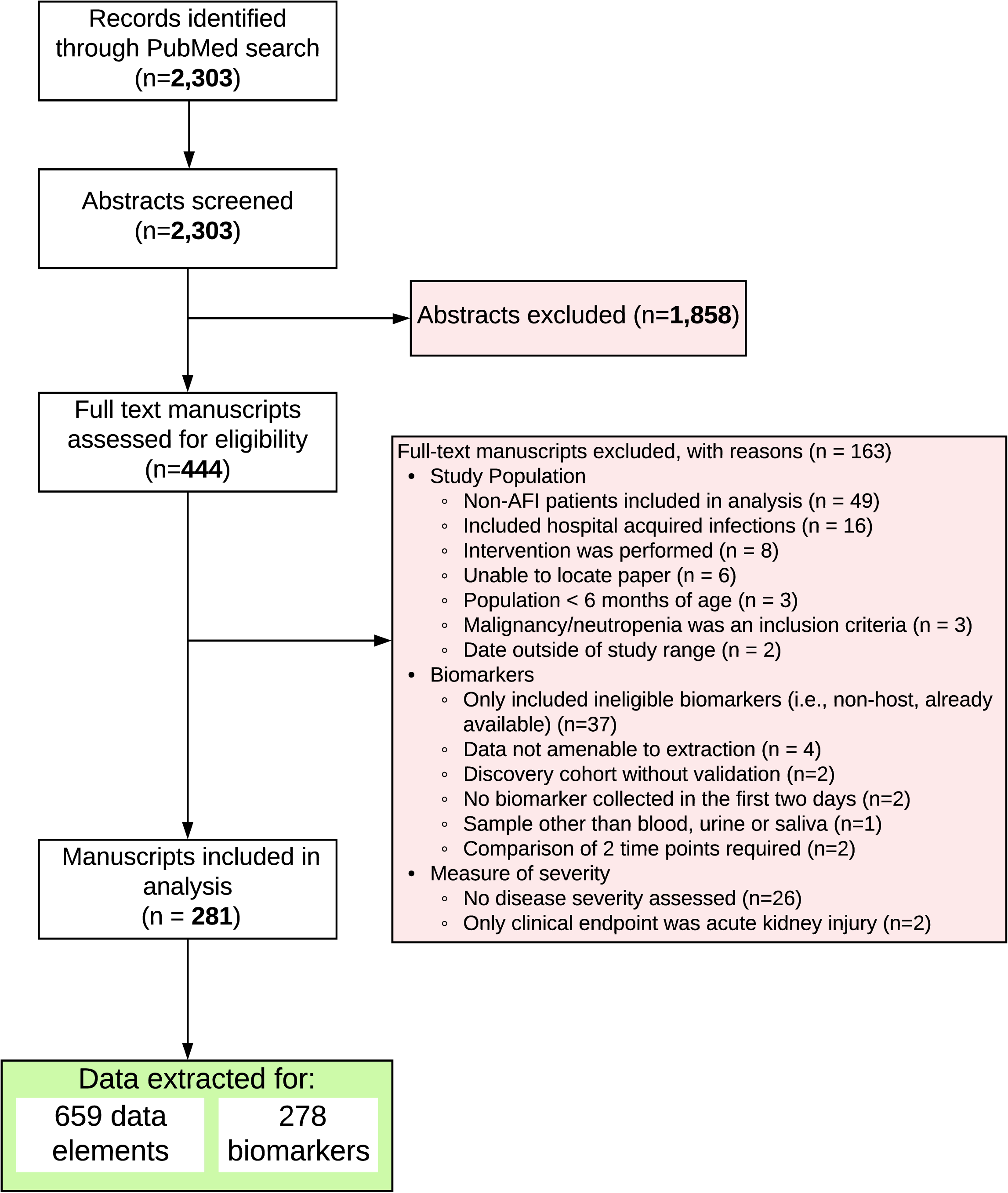
Identification, screening, eligibility assessment, and data extraction.

### Study and patient characteristics

The included manuscripts reported results for 45,737 participants in 49 countries (Figure 2). The median number of participants per study was 104 (interquartile range [IQR] 53-186). Participants were classified as having 21 distinct diagnoses or clinical syndromes (Figure 3); sepsis was the most common. Among the included manuscripts, 146 (52%) studies were conducted in 24 LMICs and included 19,513 (43%) participants. Further restricting to studies conducted in LMICs in non-ICU settings, there were 11,630 participants included in 20 countries.

**Figure 2:**
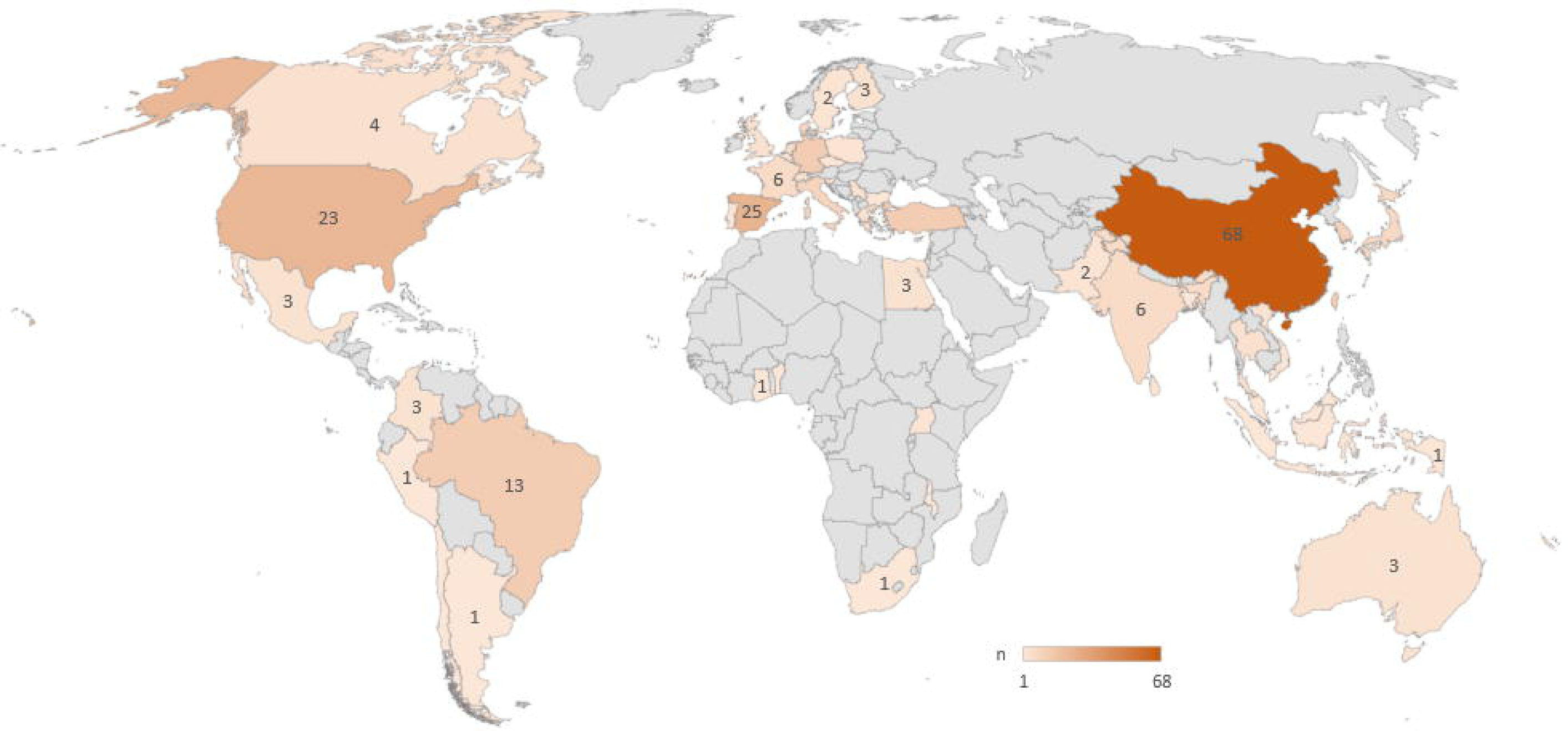
Number of manuscripts included by country.

**Figure 3:**
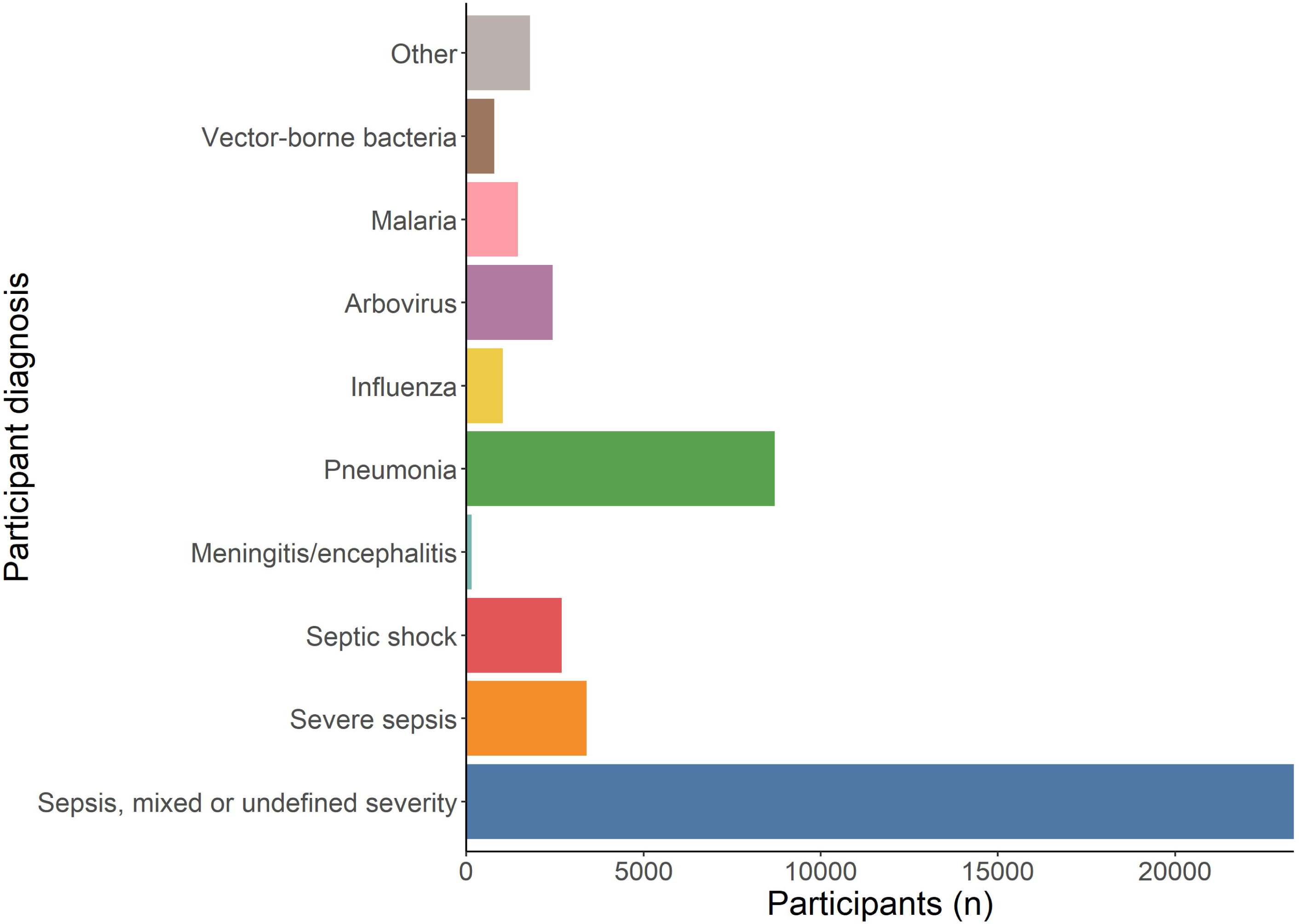
Diagnoses of participants, N = 45,737.

### Diversity of evaluated biomarkers

Data for 278 unique biomarkers were extracted. Individual publications reported a median of 1 (IQR 1-3) biomarkers. The most frequently evaluated host biomarkers were IL-6 (n = 50), IL-10 (n = 33), and TNF-α (n = 26). Most biomarkers (n = 189) were only evaluated in a single publication. There were 89 biomarkers evaluated in at least 2 studies (Supplementary Table); 52 biomarkers were evaluated in ≥ 3 studies. The most commonly studied biomarker categories were pro-inflammatory cytokines (19 biomarkers) followed by biomarkers involved in innate effector mechanisms (13 biomarkers), immune regulation (12 biomarkers), leukocyte migration (12 biomarkers) and vasoactivity (12 biomarkers) (Supplementary Figure 1).

### Biomarker performance

Mortality was the reported outcome in 193 (65%) studies. Disease-specific measures of severity were the second most commonly reported outcome (n = 53, 18%). A measure of strength of association was reported by 168 (60%) studies. The most commonly reported measure of strength of association was AUC-ROC, which was reported in 151 (54%) of studies (Table 2). Among the biomarkers studied, there were 15 biomarkers that showed a weighted mean AUC > 0.7 studied in > 500 patients over ≥ 2 studies in which ≥ 75% of studies showed an association of the biomarker with the severity outcome; 6 of which showed a weighted mean AUC > 0.75 (Figure 4). Among studies conducted in LMICs without ICU patients, there were 4 biomarkers which showed a weighted mean AUC > 0.7 studied in > 500 patients over ≥ 2 studies (Supplementary Figure 2).

**Table 2:**
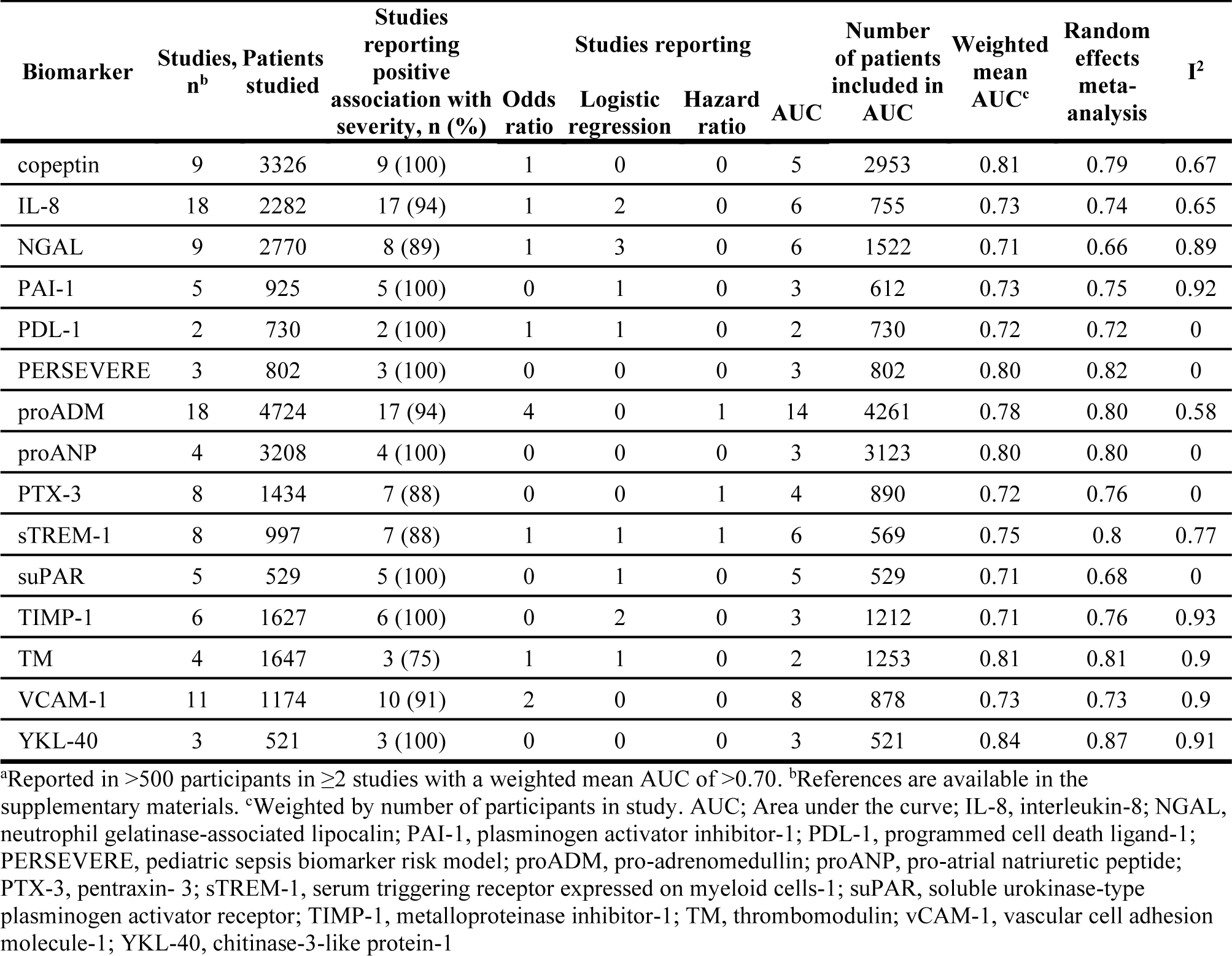
Evidence base and performance of leading^a^ biomarkers to predict acute febrile illness severity

**Figure 4:**
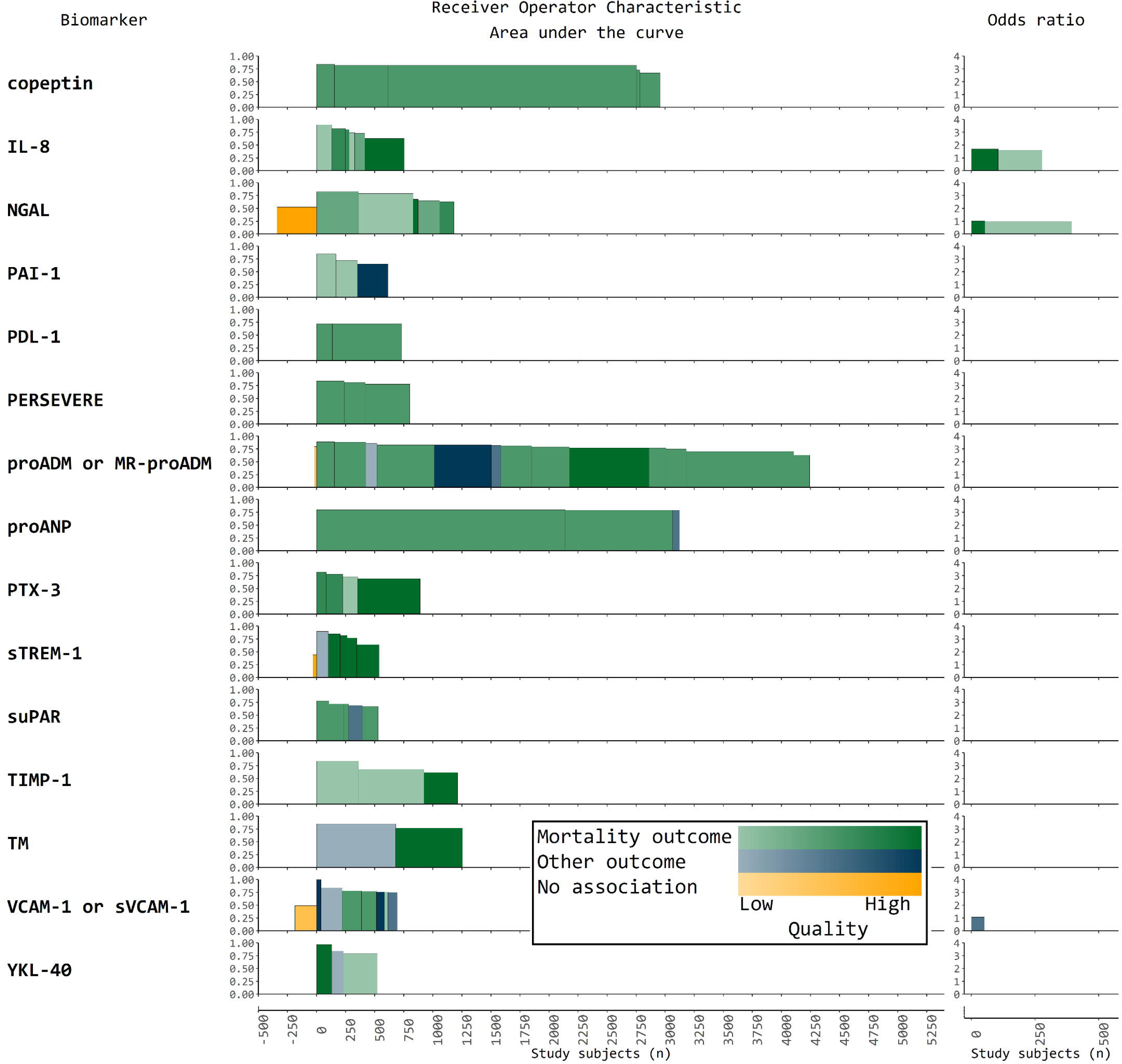
Participants studied, effect size, and quality of leading biomarkers to predict the severity of acute febrile illness. Each shaded tile represents one study. The width of the tile is proportional to the number of subjects enrolled; the height is proportional to the effect size of the association as measured by AUC or odds ratio.

## Discussion

In this scoping review, we report data for 278 novel biomarkers studied in 45,737 patients with 21 causes of acute febrile illness in 281 studies from 50 countries, yielding an understanding of the current evidence base for biomarkers to predict the severity of AFI. Among the biomarkers studied, there were 6 biomarkers that showed a weighted mean AUC greater than 0.75 in more than 500 patients over more than 2 studies. The current evidence base reports mostly the performance of biomarkers predominantly in high-income intensive care settings; a smaller, yet still robust evidence base describes the performance of biomarkers outside of ICUs in LMICs. The overall performance of biomarkers suggests that although they may aid in determining the prognosis of AFI; they are insufficient as the sole predictors of severity of febrile illness.

Biomarkers in 8 categories demonstrated potentially useful performance in multiple studies exceeding 600 total study patients, suggesting their promise for future study. The number of studies exploring a biomarker, the number of participants in each study, as well as the variation in ages, disease states, and settings in which the patient population has been studied, contributes to the breadth of evidence supporting biomarker performance. As AUC is a single number that accounts for both sensitivity and specificity independent of a cutoff value, we chose to use AUC as the measure to aggregate biomarker performance across studies. We found several biomarkers including proadrenomedullin and copeptin that had sufficiently large evidence bases and favorable performance to suggest that they may warrant further exploration as fever triage candidates. Proadrenomedullin has been studied in >4,800 patients in 18 studies across varied geographical settings, age groups, and disease states. Almost all of the studies of proadrenomedullin reported an AUC, with a weighted mean AUC of 0.78. Similarly, copeptin has been studied in >3,000 patients with a weighted mean AUC of 0.81.

Current research reporting the performance of biomarkers to predict AFI severity has not been primarily conducted in the settings where their potential impact is greatest. The highest burden of AFI is in tropical LMICs^1^ where inpatient and critical care beds are limited. The most likely intended use for a fever triage test would be among patients under assessment for the appropriate level care where the capacity for clinical assessment is limited. Most of the studies in this review were conducted in ICU settings where patients already likely show frank clinical signs of the requirement for a higher level of care. Almost half of the included studies were conducted in the US and Europe which may not represent the population and healthcare systems that would most benefit from an AFI severity biomarker.

Readily available clinical signs such as respiratory rate and simple scoring systems such as qSOFA are already used routinely to asess AFI clinical severity. The evidence compiled in this scoping review does not suggest that single biomarkers, even the most promising ones, perform sufficiently well enough to clearly supercede existing clinical severity assessment tools such as qSOFA, which has an AUC of 0.66-0.85 to predict severe or mortal AFI.[57, 58] However, unlike subjective clinical assessment, biomarkers may be standardized and not rely on the skill of a clinical assessor. Earlier experience with combining qSOFA and biomarkers have shown promising results, with a higher AUC for the combination than any individual method of prognosis assessment.[59-61]

There are a several limitations to this scoping review. The available data are highly heterogeneous in patient population, disease state, and study design. The majority of observations in this scoping review only reported the presence or absence of an association with an unfavorable outcome; only 48% of observations reported the strength of the association. Only studies published in the last 5 years were analyzed which may miss earlier and important contributions. The quality assessment that we performed was basic. Because we required only that patients be suspected or diagnosed to have a syndrome that is known to cause AFI, a significant proportion of participants may not have been febrile at the time of study enrollment which may limit the generalizability of the findings to the intended use case of evaluating febrile patients.

Biomarkers which demonstrated promising performance but have a limited evidence base such as serum Triggering Receptor Expressed on Myeloid cells-1 may show further promise as triage candidates when studied in more patients. A formal meta-analysis of pro-inflammatory cytokines and vasoactive biomarkers, or a prospective study in larger and more varied cohorts would provide higher confidence evidence than obtained in this scoping review. Additionally, it would allow the formulation and testing of algorithms with overall improved performance characteristics. Finally, prospective evaluation of biomarkers in clinical practice will be necessary to show that determination of AFI prognosis impacts meaningful treatment outcomes.

AFI impacts a significant portion of the global population every year. This scoping review synthesized a large amount of data to shortlist biomarkers that may be useful in the assessment of AFI severity as triage test candidates. Further research is necessary to determine whether biomarkers can be integrated with existing fever assessment guidelines to improve clinical practice and patient outcome.

## Data Availability

Data referenced in this manuscript is available in the supplementary materials.

## Acknowledgments

We are grateful to Rahul Rajkumar for assistance in data abstraction. We further would like to thank Tomas Jansen, Teri Roberts and Arlene Chua from Médecins Sans Frontières for their input in study design. There are no reported conflicts of interest.

